# Improvements in diarrheal disease prevalence with point-of-use water filter implementation in the informal settlement of Kibera, Kenya

**DOI:** 10.1101/2023.03.03.23286740

**Authors:** Nathan L. Tintle, Jason Westra, Kristin Van De Griend, Virginia Beard, Benjamin N. Turner, Natalie L.H. Huisman, Nicholas Dawson, Lillian Droscha, Clay Ihle, Matthew Moore, Marilyn Orellana, Luke Schutter, Lydia Snyder, Devin White, Makayla Wilson, Grace K. Goszkowicz, Brent P. Krueger, Aaron A. Best

## Abstract

**Background:** There is increasing evidence of the efficacy of point-of-use water filters on diarrhea prevalence in numerous global settings, in both observational studies and randomized experiments. Most studies, however, are focused on rural locations. Methods We use self-report household surveys to monitor a set of approximately 10,000 households receiving point-of-use water filters and WASH training in Kibera, Kenya. Twenty-five drinking water sources throughout the 7 neighborhoods were also selected for testing of E. coli, total coliform, bacterial 16S rRNA community sequencing and metals. Albendazole was provided to all households at distribution as part of the standard filter distribution protocol, with a subset of 2,642 households not receiving Albendazole at distribution, instead receiving it at the second follow-up (approximately 5 weeks after filter distribution).

**Results:** After data cleaning, a sample of 6,795 households were analyzed using mixed effects generalized linear models to account for repeated household measurements, geospatial and temporal effects, interviewer and other household covariates. Models predicted self-reported, 2-week prevalence of diarrhea. After accounting for confounding factors, self-reported diarrhea rates dropped from 52.7% to 2.2% after approximately 70 days of filter use. Field testing characterized most water sources (18 out of 25) as unsafe for Total coliforms, many for E. coli (6 out of 25), and one source above WHO health guidelines for arsenic. There was no evidence of a difference in self-reported diarrhea prevalence between households receiving Albendazole at distribution vs. those that didn’t (p>0.05).

**Conclusions:** The introduction of Sawyer filters to households in a densely populated informal settlement reduced diarrhea and other health related problems. Representative water quality testing indicates a high frequency of drinking water source contamination with E. coli and Total Coliforms but a very low frequency of dissolved metals present, above WHO guidelines for drinking water. Anti-parasitic medication distribution had little to no impact on the results. Future randomized controlled studies with objective health measures are needed to ensure cause-effect impact of the filters, and study of filter longevity in the field continues to be a critical need.

## Background

Globally, diarrhea was associated with an estimated 1.45 million deaths (1). More than 2 billion people use feces-contaminated drinking water sources, and nearly half a million deaths annually are associated with microbial contamination in drinking water (2). According to the World Health Organization (WHO), there are 1.7 billion cases of childhood diarrheal disease annually and 525,000 children under 5 die from the disease each year, making diarrheal disease the second leading cause of death in children under 5 (3). Since a major source of diarrhea is fecal pathogens via fecal-oral transmission (4,5) these lives could have been saved through clean drinking water (2) and proper hand hygiene (5,6).

The global development community invests billions of dollars each year in interventions to promote access to adequate clean water around the world (7), though only 45% of countries are on target to reach their national drinking-water targets (8). Despite global spending, interventions often underperform their targeted goals or are unsustainable, with 30–50% of water, sanitation, and hygiene (WASH) projects failing after 2–5 years (9). Clasen et al. (10) found that, while interventions are generally successful in preventing diarrhea, there is significant heterogeneity across studies.

The most often implemented water interventions in communities include Point-of-Use (POU) filters placed in households and Point-of-Collection (POC) or Point-of-Entry (POE) filters. POC/POE filters are not as effective as POU filters due to inconsistency in the availability of needed reliable supply chains and appropriate community level management (10,11). One POU option is the Sawyer PointONE® water filter. Laboratory tests with the Sawyer PointONE® water filter suggest it aligns with the United States Environmental Protection Agency standard for bacteria and protozoa removal (12). Prior studies have identified a significant decrease in diarrhea prevalence (12,13). Some studies have argued that filters in the field have been fouled and under-utilized in practice (14) (15), however, others have noted numerous limitations of these studies (16) and reasonably good performance at removing *E. coli* and coliforms over a one- to three-year period in the field (17). Thus, there is a continued need for carefully designed field trials to evaluate the efficacy of Sawyer PointONE® and other hollow fiber membrane filters. Nearly all prior studies have focused on the Sawyer PointONE® and other similar filters in rural contexts. There is a need for efficacy evaluation in urban contexts.

Currently, efforts are underway to bring Sawyer PointONE® filters to each household in the approximately 250,000-person informal settlement of Kibera, located within Nairobi, Kenya. Kibera is the largest such settlement in Africa (18). The following is a 90-day prospective analysis of self-reported diarrhea with parallel field and laboratory-based source water tests on approximately 10,000 households in Kibera.

## Methods

### Data collection

Data were collected as part of a larger campaign to install Sawyer International Bucket System® (19) water filters throughout an approximately 250,000-person informal settlement (Kibera), located within Nairobi, Kenya. Interviewers were trained by The Bucket Ministry (TBM) in conjunction with Aquora Research and Consulting, LLC (Aquora) on data collection procedures. Aquora researchers participated in onsite training in Kibera in November 2021, with a follow-up visit in March 2022 to ensure compliance with field protocols. Visits included embedding with randomly selected interviewers, as well as with interviewers identified with aberrant survey data, to improve compliance. Neighborhoods were canvased by TBM and community members were invited to distribution events of approximately 500 households (range: 60-1050). At distribution events, all households were provided a filter for their home (one per household), baseline survey data were collected, group trainings were conducted on proper filter use and basic hand washing, and individuals receiving filters were required to demonstrate filter cleaning via backflushing. Data collectors visited each home with scheduled visits at 7-10 days, 30-45 days and 60-90 days after filter distribution. To build trust, the same data collector was scheduled to conduct all three follow-up visits with assigned households. Over 100 data collectors were active in the field throughout the course of the study. Data collection procedures were approved by the Institutional Review Board at Dordt University (a federally compliant IRB: FWA00028884), by contract with Aquora.

### Survey

All household residents 18 years of age and older were eligible to participate and informed consent was received prior to participation. Receiving a filter was not contingent upon participating in the survey. A single household resident answered questions on behalf of the entire household. The surveys (baseline and follow-up) were developed by Aquora to maximize scientific rigor (valid and reliable measures), simplicity and minimize time in the field. Multiple rounds of input from stakeholders yielded preliminary surveys which were then forward and back-translated into Swahili/English by two separate leaders of the field data collection teams. Back-translation yielded minor translation inconsistencies which were then addressed with the translators yielding two reliably translated surveys. Initial training on the surveys and pilot testing in mid-November 2021 identified additional minor survey implementation questions which were addressed in the field before roll-out of the final version of the survey in late 2021.

### Survey contents and measures

Surveys were developed to address the following: basic household demographics (e.g., gender, age, number of household members, job status, school status), where household water is typically obtained, how much money is typically spent on water, self-reported diarrhea (two-weeks), self-reported impact of diarrhea on school/work participation (two weeks), medical costs (previous month), self-reported other health issues (two weeks) and self-reported hand washing. Follow-up surveys also included measures of filter utilization (i.e., how often filter buckets are refilled) and interviewer perceptions of filter utilization/functionality/user ability to clean the filter. At the time of follow-up surveys, if interviewers perceived a lack of knowledge or understanding of filter utilization and/or hand washing among respondents, interviewers retrained the respondents. In addition to the previous questions, as part of TBM organizational practices, a separate set of faith-based questions were asked of respondents after all other questions, but before the interviewer left the home. Respondents were informed that responses to these separate questions in no way impacted their eligibility to participate in the filter program.

### Sample

The target sample was 10,000 households with data collection beginning in late November, 2021 after in-field training and quality assurance were completed. Data collection ended July 3, 2022 after the last of the initial 10,000 households completed their third follow-up. Data was collected in 7 neighborhoods in Kibera. By design, this report covers the initial 10,000 households, though data collection continues across the remaining ∼90,000 households as part of the Kibera-wide implementation project being conducted by TBM.

### Water testing

Water source samples were collected in the seven neighborhoods of Kibera that were included in distribution and follow-up surveys as part of this study and were selected to represent typical drinking water sources that are used daily in Kibera. In addition, ad hoc testing of the river that runs through Kibera and other sources that could be used as drinking water was conducted, though, to our knowledge, these are not currently used as drinking water sources. For each water source tested, three, ∼20L samples were collected in portable containers after rinsing with source water. Tandem filter kits were used to test each water source. The kits included 1) one Aquagenx CBT EC + TC (Compartment Bag Test) Most Probable Number (MPN) kit (20) for in-field *E. coli* and Total Coliform testing, 2) three tandem filter assemblies for collection of full bacterial communities for 16S rRNA sequencing, and 3) three metal testing kits for collection of water for ICP-MS analysis at a certified testing laboratory in the United States. These tandem filter kits are designed to test three, independent replicates of each water source, with the exception of the single in-field EC + TC test. Tests were conducted by Aquora personnel or local TBM team members who were trained in the field by Aquora personnel according to protocols developed specifically for this testing approach (21). Information about each water source and testing procedure, including EC + TC testing results, were recorded using FastField Forms or ArcGIS Survey123® mobile device apps in real time. All testing forms were developed by Aquora. Tandem filter kits used for water sources were transported back to the United States either in airline luggage or shipped via FedEx. Upon receipt in the laboratory, tandem filter assemblies were stored at -80°C until processing; metals testing kits were acidified and submitted to ALS Environmental, Holland, MI for testing of Arsenic, Barium, Cadmium, Chromium, Copper, Lead, Nickel and Selenium. Metals results were compared to WHO guidelines for metal concentration limits in drinking water. Test results for EC + TC were analyzed to verify accuracy of in-field test interpretation by comparison of recorded results to photographs of the test kits. These validated results were translated to MPN concentrations (cells/100 mL) and assigned to WHO Health Risk Categories based on MPN. Tandem filter assemblies were processed to recover bacteria filtered from source water by individually backflushing A filters (capturing source water bacteria) and B filters (control for each replicate). Total genomic DNA was extracted from backflushed material for downstream sequencing of 16S rRNA bacterial genes. Library preparation and QA/QC were performed according to the Schloss Lab MiSeq Wet Lab SOP (22) and sequenced using Illumina MiSeq 2×250 bp v2 paired end sequencing according to manufacturer instructions. Data processing of 16S rRNA sequenced libraries was conducted on a high-performance computing cluster using the Schloss Lab MiSeq SOP mothur (23). Data analyses were conducted using custom R (24) scripts to assess bacterial community profiles and determine presence/absence of 18 bacterial genera known to contain waterborne pathogen species (*Acinetobacter, Aeromonas, Burkholderia, Campylobacter, Enterobacter, Escherichia/Shigella, Francisella, Helicobacter, Klebsiella, Legionella, Leptospira, Mycobacterium, Pseudomonas, Salmonella, Staphylococcus, Tsukamurella, Vibrio*, and *Yersinia*) (25). Sequencing data are publicly available via the NCBI Short Read Archive (SRA) under project accession number PRJN939352 (https://www.ncbi.nlm.nih.gov/bioproject/939352). In addition, custom R scripts were implemented to summarize EC + TC data, metals data, and water source metadata. Individual water quality dashboards were produced for each water source, summarizing the three data types and general water source descriptions captured through in-field surveys.

### Data cleaning

Despite numerous internal validation checks built into the survey instrument, and automatic scheduling of follow-ups, substantial data cleaning was necessary to yield a high-quality dataset ready for analysis. First, early in the project, data heterogeneity by interviewer was observed. Notably, response patterns by interviewer suggested that interviewers were not consistently implementing the field protocols. Aquora coordinated with TBM and field managers in Kibera to generate lists of interviewers who the data suggested were not consistently implementing field protocols. These interviewers were then observed in the field and received retraining. This process continued for the remainder of the project.

Second, early in the project, surveyor data suggested that data collection tablets and/or login IDs were shared between interviewers, which made tracking and identifying surveyors challenging. Again, retraining occurred for quality improvement. To address both data limitations, data from collectors who generated health outcomes in the top 5% or bottom 5% prevalence (e.g., very high or very low levels of reported health outcomes) were eliminated from the dataset prior to any of the following analysis.

Third, despite a scheduling system, there were substantial differences in the time at which surveys took place. Ideally, follow-up surveys took place at 7-10 days, 30-45 days, and 60-90 days, however, there were substantial deviations from this ideal distribution. To address this issue and ensure meaningful interpretation of results, we emphasize *days since baseline* instead of follow-up number, for most of the graphs/statistical models utilized in this report.

Finally, there were numerous other observed data inconsistencies, including (a) varying household sizes reported over time, (b) very large household sizes reported (e.g., 30+), (c) unclear survey responses, (d) surveys with substantial amounts of missing data, (e) locations mapped more than 5 miles from Kibera, (f) duplicate filter IDs, (g) extreme values (more than 4SD) for work or school missed, (h) follow-up dates recorded before distribution dates, and (i) surveys with interviewer notes that suggested poor data quality. Surveys with poor quality and/or high levels of missing data were eliminated from subsequent analyses.

To ensure that households being tracked were consistent over the course of this longitudinal study, data from households were only included for analysis if data were available at both distribution and at least one follow-up (e.g., households for which distribution data was deemed inaccurate were dropped from both distribution and follow-up analyses). After data cleaning, the total number of surveys available for analysis was 25,208, with 6,795 distribution surveys and 18,413 follow-up surveys ranging from 1 to 203 days post-distribution (see Table 1).

**Table 1.** Data available for analysis after data cleaning to improve data quality and validity.

| Time point | Distribution | Follow-up #1 | Follow-up #2 | Follow-up #3 | Total surveys |
| --- | --- | --- | --- | --- | --- |
| Surveys available | 6,795 | 6,255 | 6,153 | 6,005 | 25,208 |

### Anti-parasitic medication substudy

A small substudy was designed to attempt to isolate filter and WASH effects from the distribution of anti-parasitic medication on self-reported health. To ensure sufficient statistical power, ensure ethical treatment of participants and feasibility for field teams, while accomplishing the overall study goals, we determined that anti-parasitic medication would not be distributed at baseline or the first follow-up for distributions over a one-month period. Instead, anti-parasitic medication was distributed at the second follow-up visit for this subset of households.

In early March 2022, U.S. based team members traveled to Kibera to train data collectors/field teams on the new protocol, as well as to embed with field teams to ensure consistent implementation of all aspects of the substudy and the overall study. During March 2022, distributions to N=2642 households did not receive anti-parasitic medications until the second follow-up time point (approximately 5 weeks after filter distribution).

### Statistical methods

To correctly model the longitudinal nature of the data, and account for numerous sources of confounding, generalized, linear (or logistic) mixed effects models were used. Random effects were included in the model to account for the repeated measures nature of the data with regard to both households and interviewers (two random effect terms). Fixed effects (covariates) were included in the model to account for numerous sources of potential confounding: (a) household size, (b) seasonality (fixed effect for each month), (c) filter installed (yes/no), (d) neighborhood, (e) knowledge of WASH practices and (f) filter use. Separate models were fit for each different health and economic outcome. Similar models were fit for filter utilization, removing filter installation and filter use from the covariates list.

Results provided in this report focus on graphs of adjusted health, economic impact and utilization rates – accounting for covariates and mixed effects, and after data cleaning. Ninety-five percent confidence bands are included on all graphs. Nearly all results shown are statistically significant (p<0.001) with non-significant differences noted in the text. Covariate adjusted prevalence estimates on the graphs are obtained by selecting median levels of covariates. Figures provided use two week-categories to obtain (unsmoothed) estimates. Assessment of the impact of anti-parasitic medication was made by adding a term to the model to represent when medication was distributed.

## Results

### Sample

Table 2 provides a general overview of the 6,795 households at baseline. Most households were multi-member (average 4 members), and most had children (<18 years) living in the household (77.4%). Hand-washing was reported to be typical after defecation and before eating/food prep. All surveys were administered between November and April, in seven zones in Kibera. Follow-up surveys were completed between 2 and 16 weeks after baseline (2 weeks: 5,915 surveys; 4 weeks: 4,801 surveys; 6 weeks: 2,720 surveys; 8 weeks: 3,951 surveys; 10-16 weeks: 655 surveys).

**Table 2.** Sample Characteristics at baseline.

| Characteristics | Percent (x/N) or Mean (SD) |
| --- | --- |
| Household size (persons) | 3.99 (1.68) |
| Baseline self-report hand washing... |  |
| ...after defecation | 97.2% (6604/6795) |
| ...before eating | 98.4% (6686/6795) |
| ...before food preparation | 88.1% (5986/6795) |
| Children (<18 years) live in household | 77.4% (5259/6795) |
| Month |  |
| November | 8.5% (580/6795) |
| December | 0.3% (18/6795) |
| January | 23.2% (1576/6795) |
| February | 20.7% (1406/6795) |
| March | 33.7% (2292/6795) |
| April | 13.6% (923/6795) |
| Kibera zone |  |
| Ayany | 12.9% (878/6795) |
| Fort | 10.6% (722/6795) |
| Karanja | 6.1% (414/6795) |
| Kianda | 8.6% (587/6795) |
| Olympic | 0.4% (27/6795) |
| Raila | 43.4% (2946/6795) |
| Soweto | 18.0% (1221/6795) |

### Filter utilization and functioning

After accounting for covariates (e.g., seasonality; interviewer; see methods), approximately 97.2% of participants report having a functioning filter and using it within 6-weeks of distribution (see Fig 1). The observed increasing trend likely indicates positive impact of follow-up visits on retraining, ensuring parts are available and building confidence in the utility of the filter for participants.

**Fig. 1.**
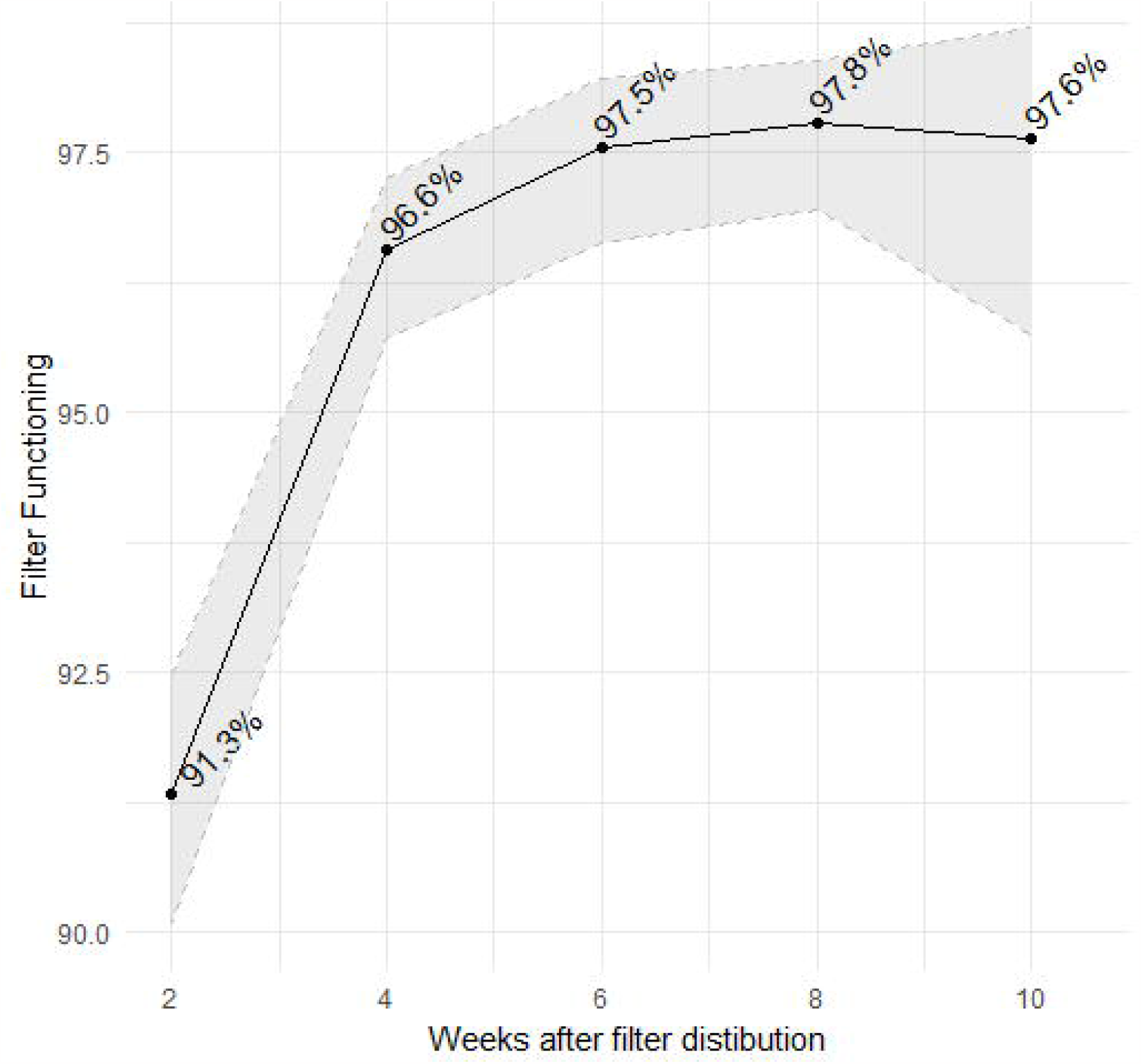
Model adjusted estimates of filter utilization^a^. ^a^ 95% confidence interval bands shown in gray

### Health impacts

Self-reported two-week diarrhea prevalence dropped substantially from baseline to 10-weeks, with the largest drop observed between 2 and 4-weeks, reaching its lowest values, and maintaining these levels, after 6 weeks. Diarrhea prevalence decreased overall (Fig 2A), and within all age brackets (Fig. 2B-D), with covariate adjusted estimates of decreased 2-week prevalence of approximately 20-fold across all ages after 6 weeks.

**Fig. 2.**
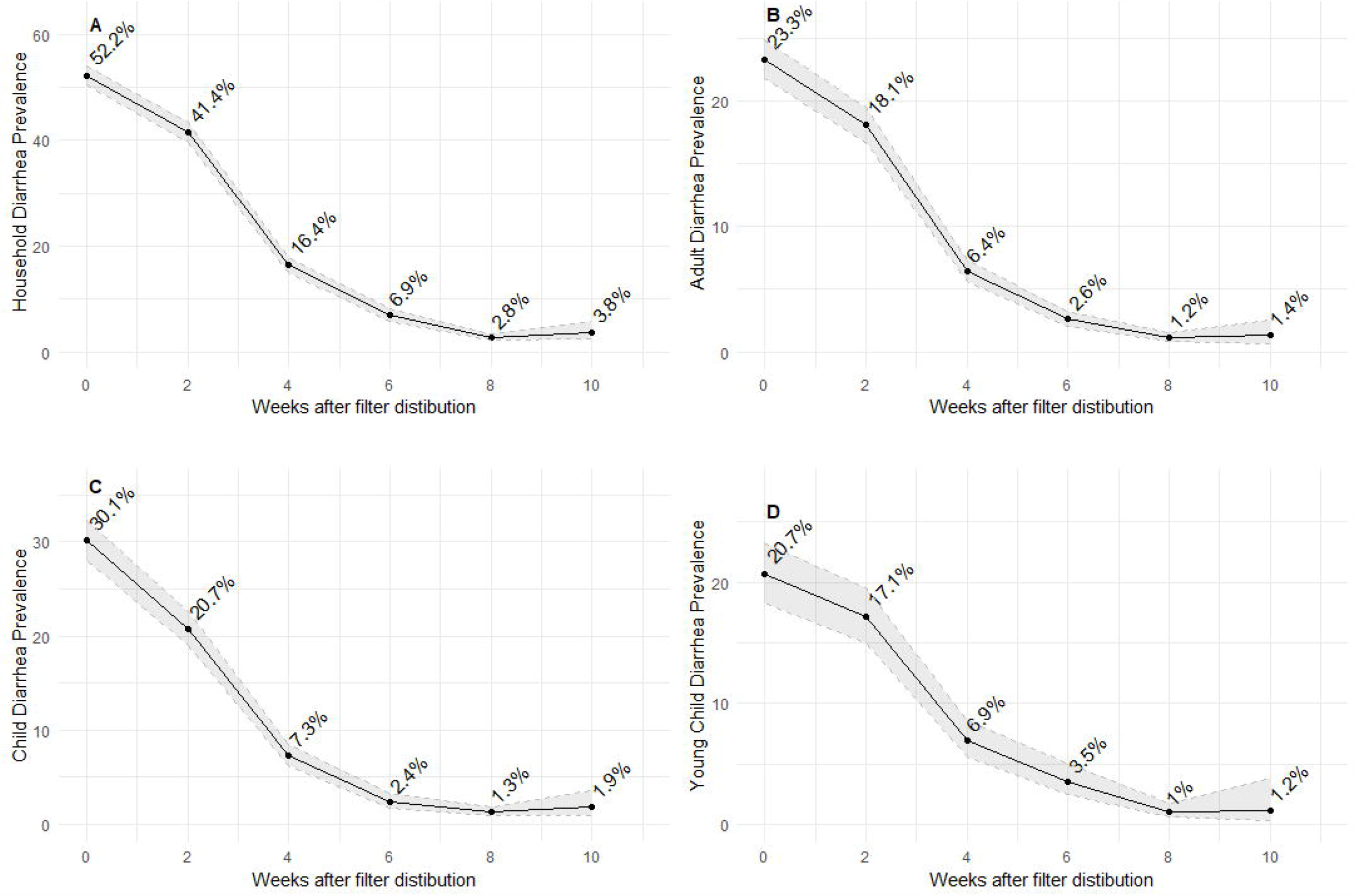
Model adjusted estimates of Diarrhea Prevalence. **A** Household diarrhea prevalence (at least one member) **B** Adult (aged 18+) diarrhea prevalence **C** School-aged child 9aged 5-17) diarrhea prevalence **D** Young child (aged 4 and under) diarrhea prevalence

Similar patterns, though with lower overall prevalence, were observed for more severe diarrhea and other reported health problems (Fig. 3).

**Fig. 3.**
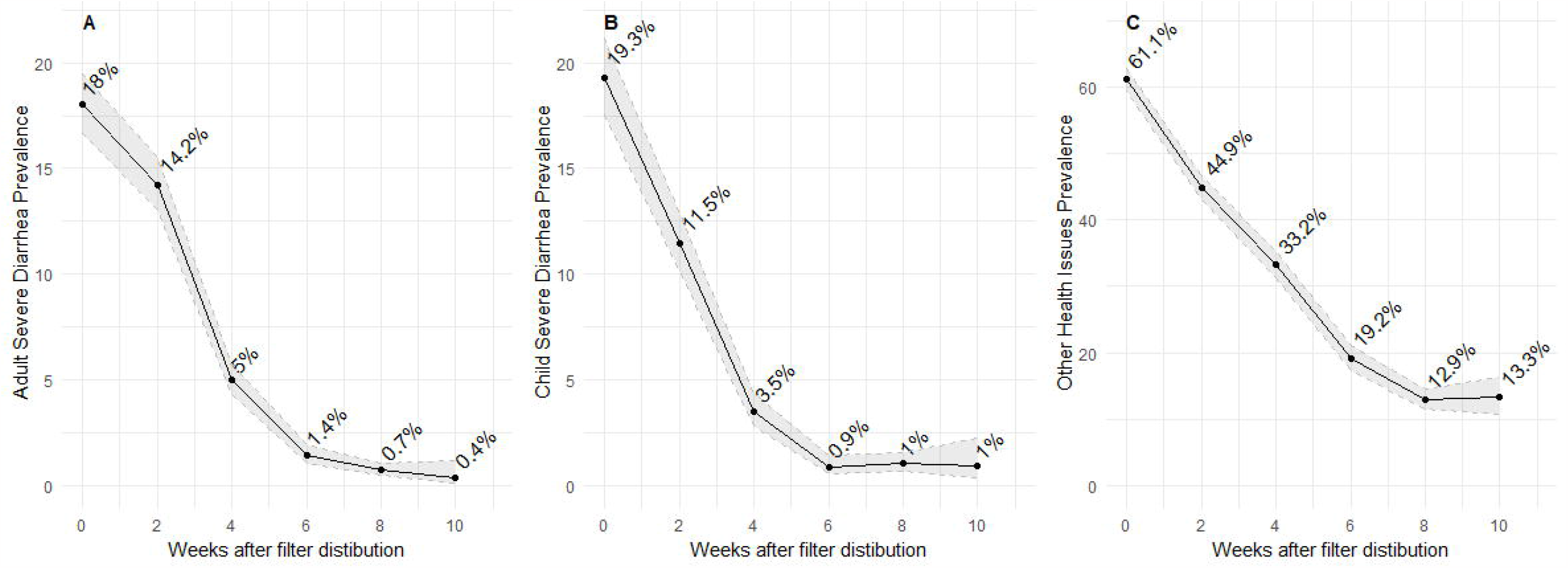
Severe Diarrhea and other health impacts. **A** Severe diarrhea (caused the individual to miss work) among adults **B** Severe diarrhea (caused the individual to miss school) among children **C** Other health issues

### Economic impact

Participants were also asked about their previous 30-day household medical and water costs. For households that previously self-reported diarrhea, medical expenses trended downward after filter distribution (Fig. 4A). There was no reduction in household water costs over time (p>0.05) (Fig. 4B). The lack of observed association between filter distribution and water costs could be explained by team member observation and documentation that drinking water in Kibera, regardless of quality, is sold from water distributors who control (illegal) taps.

**Fig. 4.**
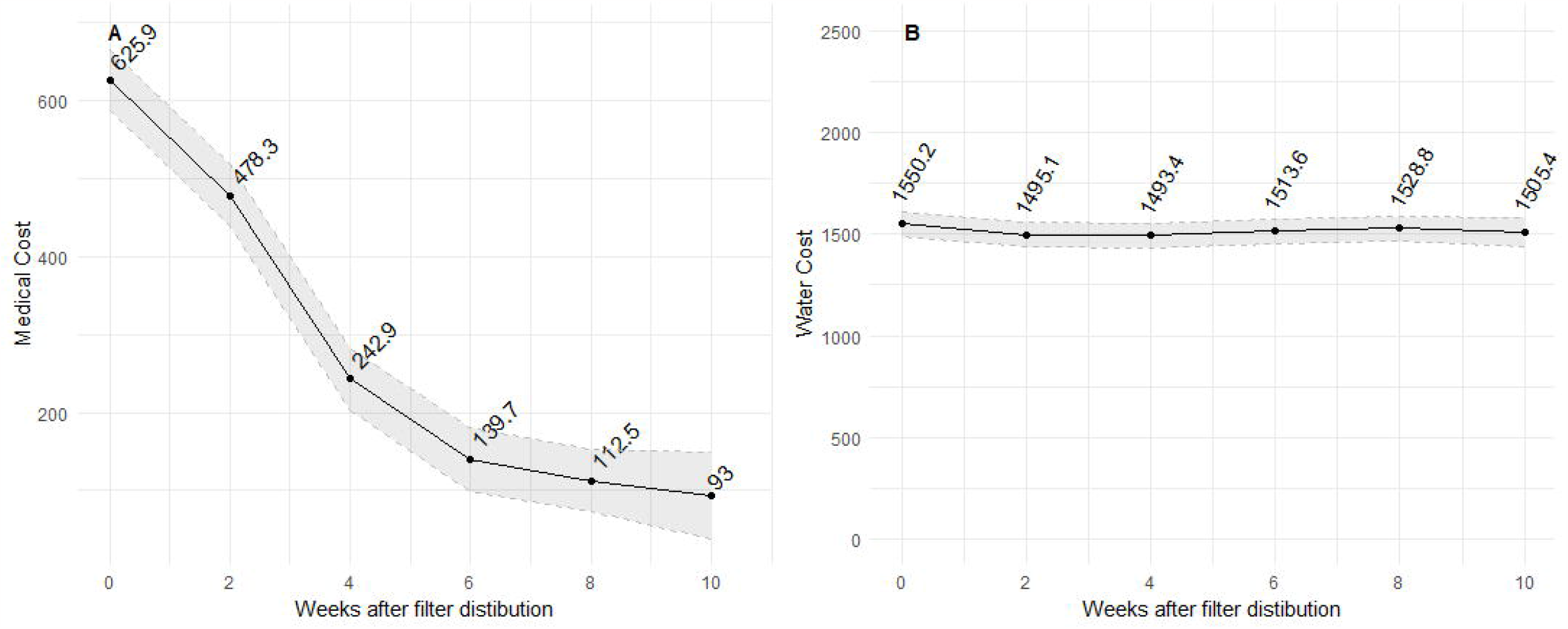
Economic Impact of intervention over time. **A** 30-day medical costs (Ksh) **B** 30-day water costs (Ksh)

### Impact of anti-parasitic medication

A subset of households were given anti-parasitic medication later than other households (see Methods). There was no statistically significant difference in self-reported diarrhea between households that were given delayed anti-parasitic at any point in the study (Fig. 5; p>0.05 at all time points and overall).

**Fig. 5.**
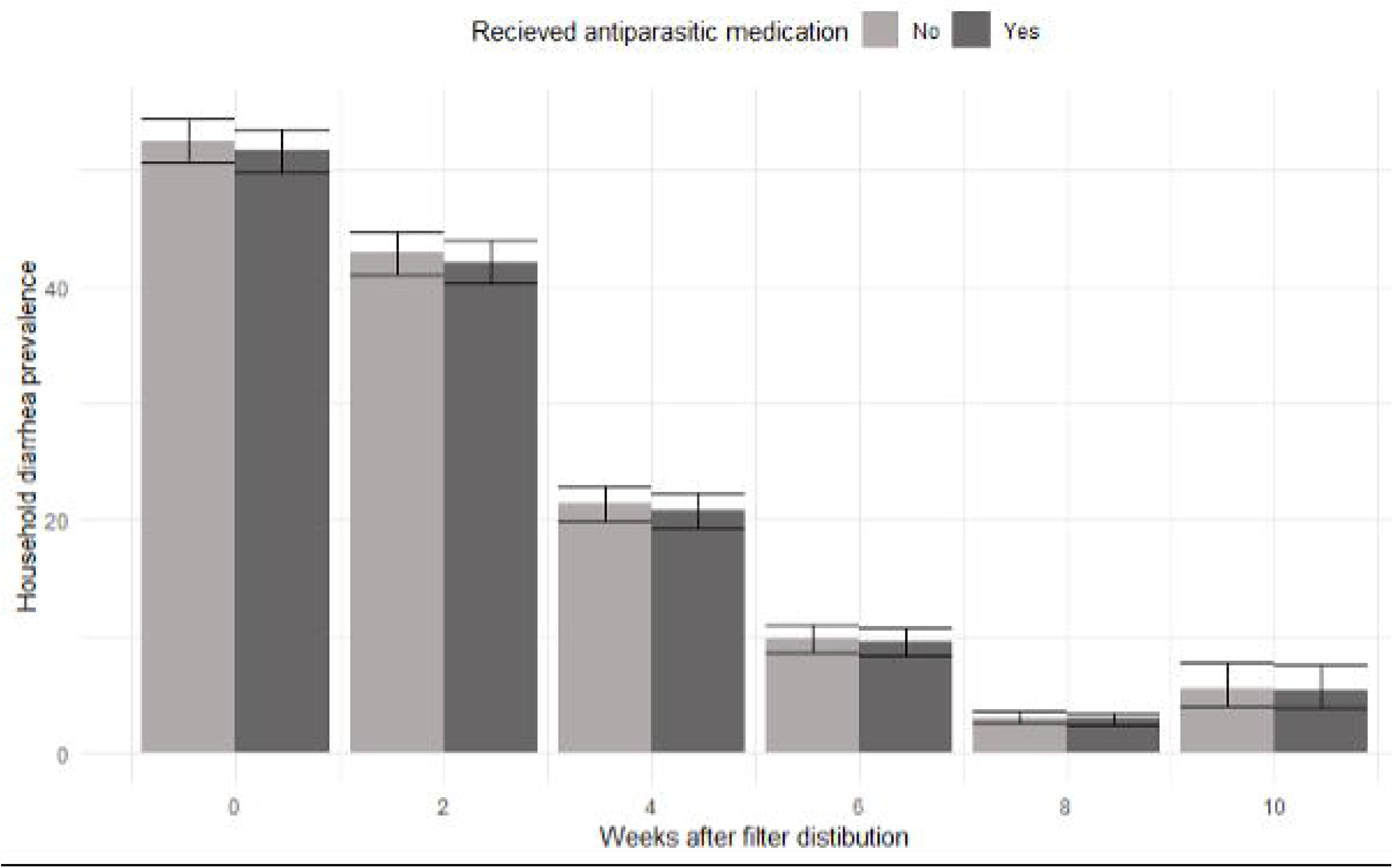
Self-reported diarrhea prevalence by anti-parasitic medication group.

### Impacts of WASH training

Attempts to isolate the effects of WASH training from filter implementation were unsuccessful, because nearly all households utilized the filters (> 97% after six weeks, see Fig. 1). Self-reported hand washing rates suggest that WASH training had a small (though not statistically significant) positive impact: individuals claimed to wash their hands after defecation (97.4% distribution; 98.3% follow-up), before eating (98.6% distribution; 98.5% follow-up) and before food prep (86.7% distribution; 92.6% follow-up).

### Water test results

At the time of filter distribution, 25 water sources in distribution neighborhoods of Kibera were tested (21 sources actively used for drinking water, 3 sources that could be used for drinking water, and 1 surface source that is not used for drinking, according to residents of Kibera). Samples were collected from the range of possible water source types for residents in these neighborhoods (Fig. 6) in November 2021, March 2022, and May 2022.

**Fig. 6.**
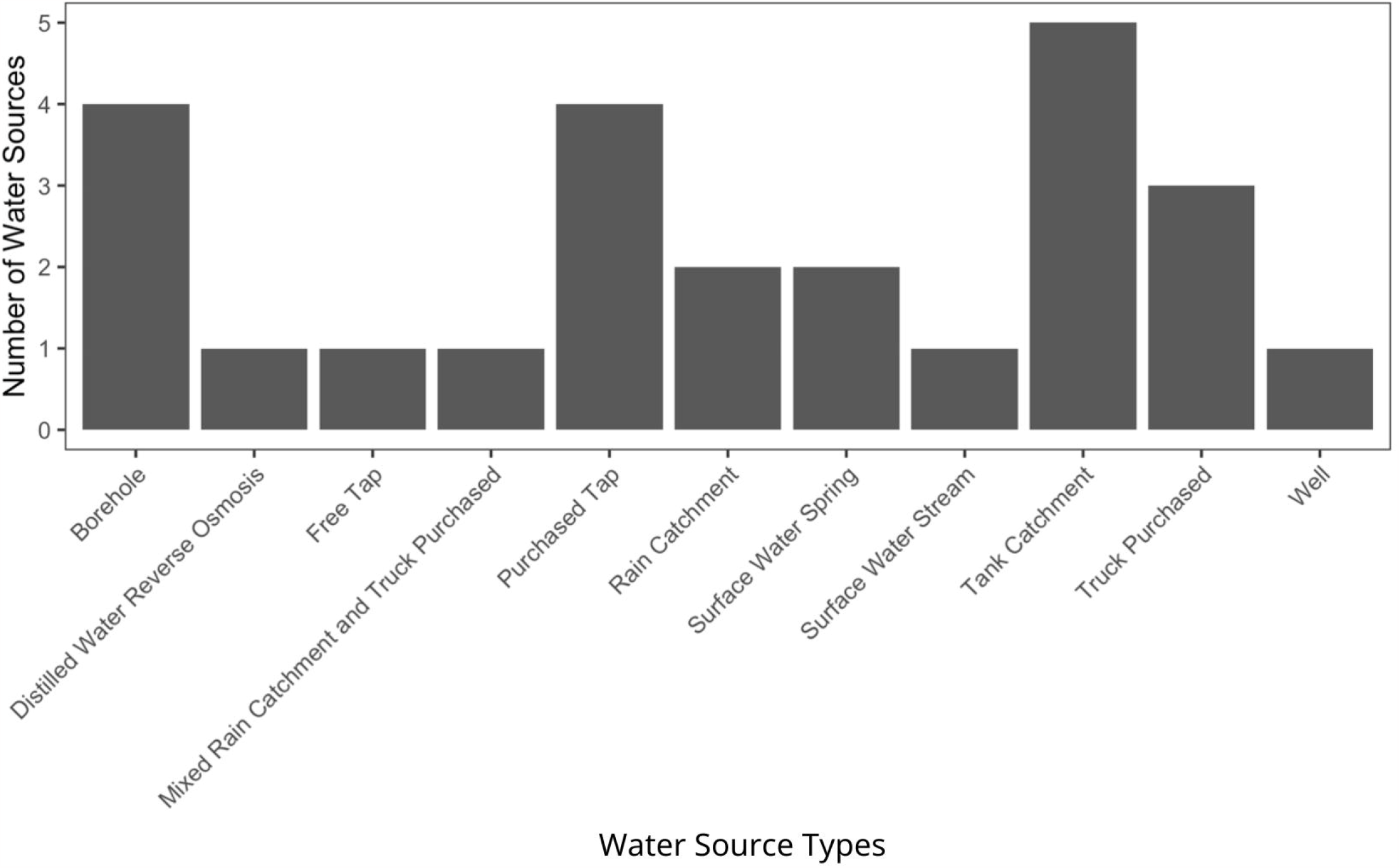
Distribution of water source types sampled in Kibera.

### Results of Bacterial Testing

Results from in-field testing of *E. coli* and Total Coliform concentrations show that most water sources available to residents of Kibera pose a high risk of water-borne illness. “Unsafe” *E. coli* levels were found in 6 water sources, according to WHO-categorization, while only 13 water sources fell into the “Low Risk/Safe” *E. coli* categories (Fig. 7A). Furthermore, 18 water sources were categorized as “Unsafe” for Total Coliforms with only 2 “Low Risk/Safe” (Fig. 7B). It is clear from these data that most water sources contain bacterial contamination at levels that are unsafe for human consumption.

**Fig. 7.**
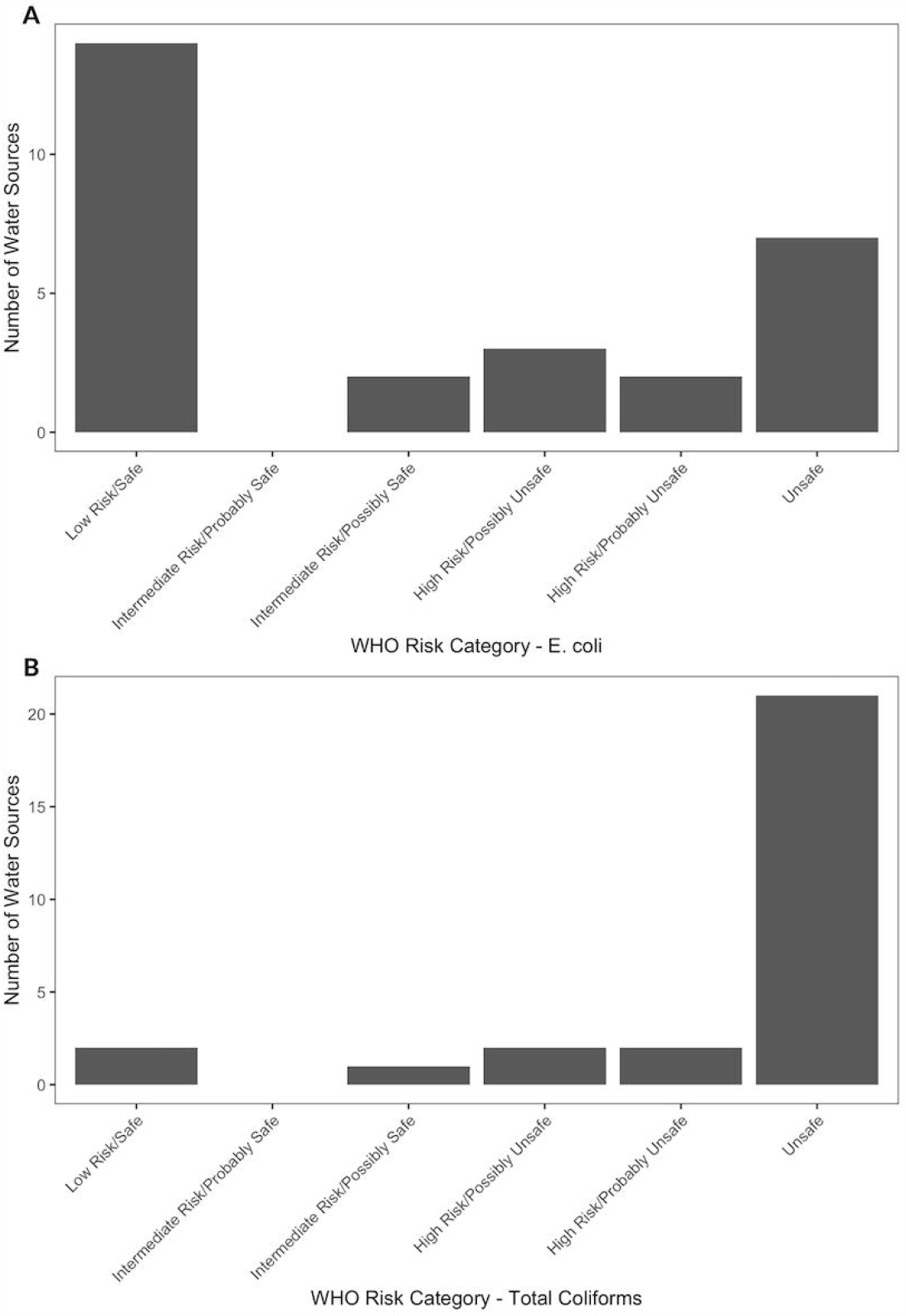
Field testing of water sources. **A** Field testing of water sources for *E. coli* concentration. Aquagenx Compartment Bag EC+TC Most Probable Number tests were conducted for all water sources sampled. Based on the determined MPN (cells/100 mL), each water source was categorized into a WHO Health Risk category. The number of water sources in each category is depicted by the height of the bar for each category. **B** Field testing of water sources for Total Coliform concentration. Aquagenx Compartment Bag EC+TC Most Probable Number tests were conducted for all water sources sampled. Based on the determined MPN (cells/100 mL), each water source was categorized into a WHO Health Risk category. The number of water sources in each category is depicted by the height of the bar for each category.

Other potential pathogens associated with drinking water were identified in all water sources tested by bacterial community sequencing (Table 3). This methodology allows for a more specific description of potential health risks than the EC+TC MPN testing methods. Each of the bacterial genera listed are known to cause either respiratory, gastrointestinal, or skin, eye and wound diseases transmitted through contaminated water. All 24 water sources with sufficient sequencing read depth were observed to contain at least three of the genera listed in the table. Water source 370, representing the river that runs through Kibera and that is not used as a drinking water source, contained seven of the genera.

**Table 3.** Presence or Absence of Potential Bacterial Pathogens^a^.

| Water Source | *Acinetobacter* | *Aeromonas* | *Campylobacter* | *Escherichia-Shigella* | *Legionella* | *Leptospira* | *Mycobacterium* | *Pseudomonas* | *Staphylococcus* | *Yersinia* |
| --- | --- | --- | --- | --- | --- | --- | --- | --- | --- | --- |
| **312** | **Yes** | **Yes** | No | No | **Yes** | No | No | **Yes** | No | No |
| **313** | No | No | No | No | **Yes** | No | No | No | **Yes** | No |
| **316** | **Yes** | **Yes** | No | **Yes** | **Yes** | No | **Yes** | **Yes** | No | No |
| **352** | **Yes** | No | No | No | **Yes** | No | No | No | **Yes** | No |
| **364** | No | **Yes** | No | **Yes** | **Yes** | No | **Yes** | No | No | No |
| **365** | No | **Yes** | No | No | **Yes** | No | No | **Yes** | No | No |
| **367** | No | No | No | No | **Yes** | No | **Yes** | **Yes** | **Yes** | No |
| **368** | No | **Yes** | No | No | **Yes** | No | **Yes** | No | No | No |
| **369** | **Yes** | **Yes** | No | No | **Yes** | No | No | **Yes** | **Yes** | **Yes** |
| **370** | No | **Yes** | No | **Yes** | **Yes** | **Yes** | **Yes** | No | **Yes** | **Yes** |
| **371** | **Yes** | No | No | No | **Yes** | No | No | **Yes** | No | No |
| **373** | No | **Yes** | **Yes** | **Yes** | No | No | No | No | No | No |
| **374** | No | No | No | No | **Yes** | No | No | No | **Yes** | No |
| **375** | No | No | No | No | **Yes** | No | **Yes** | **Yes** | **Yes** | No |
| **377** | No | No | No | **Yes** | No | No | No | **Yes** | **Yes** | No |
| **389** | **Yes** | **Yes** | No | No | No | No | **Yes** | No | **Yes** | No |
| **391** | **Yes** | **Yes** | No | No | **Yes** | No | No | **Yes** | **Yes** | No |
| **392** | No | **Yes** | No | No | **Yes** | No | **Yes** | No | No | No |
| **394** | **Yes** | No | No | **Yes** | **Yes** | No | **Yes** | No | No | No |
| **397** | **Yes** | **Yes** | No | No | **Yes** | No | No | No | No | No |
| **401** | **Yes** | **Yes** | No | No | No | No | No | **Yes** | No | No |
| **402** | **Yes** | **Yes** | No | **Yes** | **Yes** | No | **Yes** | **Yes** | No | Yes |
| **403** | **Yes** | No | No | No | **Yes** | No | **Yes** | **Yes** | No | No |
| **404** | **Yes** | **Yes** | No | No | No | No | No | **Yes** | No | No |

<sup>a</sup>16S rRNA community sequencing of 24 water sources was used to identify the presence or absence of 18 bacterial genera with known waterborne pathogenic species. Ten genera were identified amongst the samples tested. All samples contained at least three genera. Presence is indicated as “Yes” and bolding.

The results from both *E*.*coli* + Total Coliform and the bacterial community testing support the high baseline rates of diarrhea and potentially other waterborne health issues that were self-reported by participants.

### Results of Dissolved Metals Testing

To assess water quality for potential long term health effects, we tested all the sources sampled for the presence of eight metals known to cause disease and for which the WHO sets out allowable limits in drinking water. Metals testing of drinking water sources showed that all but one source had either undetectable levels or detectable levels below WHO health guidelines (Table 4) (25). One drinking water source (Source 374) showed arsenic levels above WHO health guidelines. Water source 374 is a borehole drilled near a local hospital; the water is intended for use by the hospital, but it is also available as a free source of water to the local community. It is estimated that this source serves around 70 people per day. Water source 370 (river) showed chromium levels above WHO guidelines, but this source is not used for drinking water.

**Table 4.** WHO Above/Below health guidelines for metals in drinking water^a^.

| **Water Source** | **Arsenic** | **Barium** | **Cadmium** | **Chromium** | **Copper** | **Lead** | **Nickel** | **Selenium** |
| --- | --- | --- | --- | --- | --- | --- | --- | --- |
| **312** | No | No | No | No | No | No | No | No |
| **313** | No | No | No | No | No | No | No | No |
| **316** | No | No | No | No | No | No | No | No |
| **352** | No | No | No | No | No | No | No | No |
| **364** | No | No | No | No | No | No | No | No |
| **365** | No | No | No | No | No | No | No | No |
| **367** | No | No | No | No | No | No | No | No |
| **368** | No | No | No | No | No | No | No | No |
| **369** | No | No | No | No | No | No | No | No |
| **370** | No | No | No | **Yes** | No | No | No | No |
| **371** | No | No | No | No | No | No | No | No |
| **372** | No | No | No | No | No | No | No | No |
| **373** | No | No | No | No | No | No | No | No |
| **374** | **Yes** | No | No | No | No | No | No | No |
| **375** | No | No | No | No | No | No | No | No |
| **377** | No | No | No | No | No | No | No | No |
| **389** | No | No | No | No | No | No | No | No |
| **391** | No | No | No | No | No | No | No | No |
| **392** | No | No | No | No | No | No | No | No |
| **394** | No | No | No | No | No | No | No | No |
| **397** | No | No | No | No | No | No | No | No |
| **401** | No | No | No | No | No | No | No | No |
| **402** | No | No | No | No | No | No | No | No |
| **403** | No | No | No | No | No | No | No | No |
| **404** | No | No | No | No | No | No | No | No |

<sup>a</sup>Water sources were tested for eight metals with regulatory guidelines from the WHO. Metals detected at concentrations above the WHO guidelines are indicated with “Yes” and bolding. A “No” indicates that metals were not detected at concentrations above the guidelines, however, it is possible that a metal was present but below the limits (not shown). One drinking water source (374) shows arsenic levels above the guidelines. Chromium is present in one water source (370), which is not used as drinking water source.

## Discussion

The Sawyer filter, combined with WASH training, leads to reductions in self-reported diarrhea rates, and improvement in self-reported economic measures in Kibera at rates in line with those observed in non-urban contexts in prior studies (21). Filter utilization rates are high, as observed in other contexts, though utilization increases after the first follow-up visits with recipients. This outcome suggests that the group distribution model is effective, but that the first follow-up is critical for retraining and to ensure filter utilization. Finally, there was no measurable effect of adding anti-parasitic drugs along with the filter and WASH training. Attempts to further isolate filter effects from WASH training were unsuccessful due to the large percentage of filter utilization.

### Limitations

While the study design and analysis of this study were carefully planned and implemented, there are limitations worth noting. First, this study is observational. Thus, the resulting associations are not necessarily causal. However, given (a) the prospective nature of the study and (b) the control of numerous covariates via the statistical modeling process, the resulting associations are strongly suggestive. Notably, however, randomized controlled trials are necessary to confirm the suggestive associations as truly causal. Second, this study is based on self-reports of health, economic and other outcomes. While self-reports of health outcomes are generally valid and reliable, self-report can be substantially impacted by a variety of biases (interviewer, recall, etc.). Thus, the best way to view the associated results is (a) in light of other similar studies using self-reported measures (comparatively) and (b) in terms of relative changes in self-reported outcomes (e.g., 2-fold reduction, etc.). Caution should be used if attempting to utilize the data as true measures of absolute levels of disease or economic benefit. Third, to minimize interviewer time, a single member of the household was interviewed and reported on the health of the entire household. Thus, self-reported information is further confounded by the ability of the respondent to know and recall health, economic and utilization information for all individual household members.

Efforts were made to clean the data and we are assuming that missing data are approximately distributed at random. Efforts were made to separate questions related to filter utilization, health impacts and similar questions reported here from other survey questions and related discussion with surveyors about faith-based topics. However, there is no guarantee that order effects/carry-over effects from the discipleship process run by the interviewer are not impacting study findings.

The study was implemented over a 6-month period in Kibera (Nov 2021 – May 2022). While statistical models accounted for temporal effects, the 6-month period study means that seasonal cycles of rainy/dry, which can impact water access, cleanliness and related infectious disease cycles were only observed for 6 months of a calendar year. Notably, however, the study period covered parts of two rainy seasons (Nov/Dec; Apr/May), as well as a dry season (Jan/Feb/Mar). Data were not collected during the winter dry season (Jun-Oct), which sees the coldest temperatures of the year and peak incidence of infectious diseases that typically spike in winter months (e.g., influenza).

Water quality testing was limited to 25 water sources in seven neighborhoods in Kibera. While this gives a representative sense of the types of water sources used, it was not designed to be a comprehensive study of water quality in the area. Sequencing of bacterial communities targeting the 16S rRNA gene limits identification of bacteria to the genus level. This allows for the identification of genera that are known to contain bacterial waterborne pathogens as members. However, this method is not a direct detection of pathogens and should not be interpreted as such. Water quality testing of chemical contaminants was limited to dissolved metals. Other types of chemical contaminants found in urban environments were not tested (e.g., volatile organic chemicals (VOCs) associated with petroleum and other organics).

### Implications

Overall, diarrhea reduction was similar to that observed in other contexts, suggesting the effectiveness of the Sawyer PointONE® filter in an informal, urban settlement. Furthermore, in this context, filter distributions were done in a group format, which improves the efficiency and consistency of training and data collection. However, data analysis suggests that some households (∼ 10%, see Fig. 1) are not using or receiving the filter benefits until after the first follow-up which occurs at the individual home with a single data collector who provides training. The group distribution model appears to hold promise (similar diarrhea reduction to what has been seen in other contexts), however, the need for more purposeful follow-ups in this model is worth noting and suggests further study of the economics of the group distribution model for organizations leading filter distribution efforts.

Bacteria were found in most samples, while only one drinking water source had a heavy metal concern. Overall, the results of bacteria and metals testing support the use of hollow-fiber membrane filters in Kibera and do not indicate the need for secondary level treatment to remove metals. The Sawyer PointONE® filter is designed to remove bacteria, as identified in the testing done here. These results underscore the need to conduct water source testing when choosing a filter technology, to ensure that the technology meets the needs of the sources being used for drinking water. Continued monitoring of water sources and the filters being used is required to ensure the appropriateness and effectiveness of the remediation solutions.

In this intervention, anti-parasitic medication distribution and WASH training also take place at the time of distribution, with repeated WASH training at follow-up visits as needed. Thus, health benefits should be attributed to a mix of the point of use filter, WASH training and anti-parasitic medication distribution. However, we were able to separate out the impact of anti-parasitic medication on self-reported outcomes in a substudy. The anti-parasitic medication distribution showed little to no evidence of effect. One reason for this lack of impact may be due to the limited nature of the antiparasitic medication intervention. Due to Kenyan regulatory constraints, the distribution of antiparasitic medications consisted of only a single, low dose of Albendazole at baseline for most participants in the study (the substudy pushed the timing of this dose to the second follow-up). While the anti-parasitic substudy took place entirely in March to meet feasibility concerns of TBM, potentially confounding seasonal health effects with the treatment arm, no noticeable change in self-reported prevalence of diarrhea at baseline was observed before (February) or after (April) the substudy, making it unlikely to materially contribute to the findings observed here. We also note that the substudy on anti-parasitic drug treatment was analyzed with intent-to-treat analysis, which ignores whether or not individuals chose to take the anti-parasitic drug and at what time. Finally, we note that while randomized controlled trials (RCTs) remain the gold standard to attribute causality to interventions, practical concerns (feasibility; ethical) regarding implementing an RCT for the present study led to the implementation of the study protocol outlined here. Statistical adjustment of confounding variables, a straightforward field protocol, and oversight from the Aquora team mitigate many of the potential limitations of this non-randomized interventional trial.

## Conclusions

While this study provides additional evidence of the efficacy of the Sawyer PointONE® filter in a novel context (urban informal settlement), future controlled trials are needed to isolate and attribute causality to the filter. Furthermore, additional studies are needed which further validate the use of self-reported health measures, a single respondent from the household, and the potential impact of a combined health/evangelism approach. In practice, we strongly recommend that water quality testing, including dissolved metals and other potential contaminants, is needed to ensure that intervention choice is targeted to the water issues of the household. Finally, there is a desperate need for long term studies of point of use filter utilization, filter cleaning compliance and long-term health effects. The observed length of the present study is hindered by a rather short (90-day) follow-up period. However, this lack of research on continued adoption of water intervention technologies and practices is an industry-wide problem. Without such studies that specifically focus on the long-term effects of filter systems and hygiene interventions, there is a lack of knowledge of whether or not the devices are providing consistent and reliable access to safe drinking water (26,27).

## Data Availability

Summary and/or de-identified data is available from the corresponding author upon reasonable request.

## Abbreviations

CBT EC + TC: (Compartment Bag Test)
(MPN): Most Probable Number
(PoC): Point-of-Collection
(PoE): Point-of-Entry
(PoU): Point-of-Use
(TBM): The Bucket Ministry
(WASH): Water, sanitation, and hygiene
(WHO): World Health Organization

## Declarations

### Ethics approval and consent to participate

Data collection procedures were approved by the Institutional Review Board at Dordt University (a federally compliant IRB: FWA00028884), by contract. All participants included in this manuscript consented to participate.

### Consent for publication

All participants consented for de-identified, summary data to be published.

### Authors’ contributions

NT, KDVG, BPK and AAB conceived of the study. NT and AAB drafted the bulk of the final manuscript, with GKG, KVDG and BPK providing substantive revisions. JW and NT led and implemented statistical analyses. VB and MW provided important context, data interpretation and literature review. BNT, ND, LD, MM, MO, LSc, LSn and DW processed water quality kits. BNT, CI and MM performed sequencing. NLH processed metals testing data. GKG executed revision of the manuscript. All authors read and approved the final manuscript.

## Acknowledgments

None.

